# Delivered but Discharged Without Assessment: A National Multilevel Analysis of Missed Maternal Postpartum Assessment Before Discharge After Facility Birth in Ethiopia, 2024–2025 EDHS

**DOI:** 10.64898/2026.11.16.26366337

**Authors:** Bereket Agumas Kegne, Dawit Mengesha Merhatsedk, Besheer Yeshaw Mohammed, Suleiman Ayalew Belay, Eliyas Addisu Taye

## Abstract

**Background:** Missed maternal postpartum assessment before discharge after facility birth is an under-measured indicator of intrafacility quality of care that previous Ethiopian studies did not isolate in nationally representative analyses. This gap matters because women may reach facilities for childbirth yet leave without a basic maternal clinical assessment during the highest-risk postpartum period.

**Methods:** We analyzed national 2024–2025 Ethiopia Demographic and Health Survey data for 3,645 women aged 15–49 years whose most recent live birth occurred in a health facility. Analyses incorporated sampling weights and complex survey design. Factors associated with missed assessment were examined using multilevel mixed-effects logistic regression accounting for enumeration-area clustering. Adjusted odds ratios (AORs) and 95% confidence intervals (CIs) were reported.

**Results:** Overall, 40.1% (95% CI: 37.0–43.3) of women delivering in health facilities did not receive a maternal postpartum assessment before discharge. Missed assessment was less likely following caesarean birth than vaginal delivery (AOR 0.32, 95% CI: 0.19–0.53) and among women with pregnancy complications (AOR 0.46, 95% CI: 0.34–0.63). Antenatal care showed a dose-response association, with lower odds among women with 4–7 visits (AOR 0.49, 95% CI: 0.27–0.89) and ≥8 visits (AOR 0.43, 95% CI: 0.23–0.81) compared with no ANC. Geographic inequities persisted, including higher odds in Somali (AOR 2.07, 95% CI: 1.02–4.18).

**Conclusions:** A substantial proportion of facility births in Ethiopia end without a maternal assessment before discharge, indicating that institutional delivery coverage can overestimate effective postpartum care when discharge processes are weak.

## Introduction

The postpartum period remains one of the most dangerous yet neglected phases of the maternal and newborn continuum of care, and the first 24 hours after childbirth carry the highest risk of severe morbidity and death for both women and newborns[1-3]. Global progress in reducing maternal mortality remains off track for Sustainable Development Goal 3.1, while haemorrhage, hypertensive disorders, sepsis, and other direct obstetric causes continue to drive a large share of preventable deaths[4]. Most maternal deaths from haemorrhage and sepsis occur during the postpartum period, and nearly half of all postpartum maternal deaths occur on the first day after birth[1, 4].

WHO recommendations on maternal and newborn care for a positive postpartum experience emphasize that postpartum care should extend beyond survival and contact coverage to the quality of care, including appropriate maternal and newborn assessment, discharge readiness, counselling, and continuity of care after leaving the facility[5]. Recent guideline syntheses reinforce these recommendations[6]. Yet measurement of this phase remains underdeveloped: a scoping review of postpartum care quality measures found no indicators for WHO-recommended criteria to be assessed before discharge, highlighting a major blind spot in routine monitoring[7]. For that reason, failure to assess women before discharge is better understood as a gap in intrafacility quality of care than as a simple problem of postpartum service utilization.

Evidence from low- and middle-income countries indicates that substantial gaps remain in the quality and adequacy of postpartum care despite increasing institutional delivery. Recent pooled analyses of DHS data reported that only 41.4% of women received quality maternal postpartum care, while studies from sub-Saharan Africa documented persistently low coverage of adequate postpartum care and considerable regional inequalities[8, 9]. However, these studies focused on postpartum care utilization or adequacy rather than whether women received an essential maternal assessment before discharge after facility birth.

Ethiopia similarly illustrates the gap between contact with maternity services and receipt of effective postpartum care. Despite progress in expanding maternal health services, maternal mortality remains high, with the most recent internationally comparable estimate placing Ethiopia’s maternal mortality ratio at 267 per 100,000 live births in 2020, a 72% decline since 2000, but still well above the SDG 3.1 target of 70 per 100,000 by 2030[10]. Previous national and subnational studies have consistently documented low and inequitable postpartum care utilization and identified factors such as urban residence, maternal education, household wealth, media exposure, antenatal care attendance, facility delivery, caesarean birth, and delivery complications as important determinants[6, 11]. However, these studies still primarily assessed postpartum care utilization after childbirth or within recommended postpartum timeframes rather than whether women who delivered in health facilities received an essential maternal assessment before discharge[11].

This gap is important because women may reach health facilities for childbirth yet leave without receiving a basic maternal clinical assessment during the period of greatest vulnerability following delivery[12]. Therefore, missed maternal postpartum assessment before discharge represents an important indicator of effective facility-based care rather than postpartum service utilization alone. To the best of the available evidence, this study is among the first nationally representative analyses using the 2024–2025 Ethiopia Demographic and Health Survey to examine this outcome in Ethiopia. Although household surveys cannot directly measure facility processes, factors such as staffing, workload, adherence to discharge procedures, provider accountability, and quality-improvement practices may influence whether assessments are completed after delivery[3]. By applying a multilevel framework, this study provides new evidence on individual, obstetric, facility-related, and contextual factors associated with missed maternal postpartum assessment within a nationally weighted sample[8].

Understanding this gap has important implications for evaluating maternal health progress. High institutional delivery coverage may overestimate improvements in effective coverage and continuity of care if women are discharged without essential postpartum assessment[13]. Evidence on the prevalence and predictors of missed assessment can help identify vulnerable groups, reveal points of failure within the maternal care pathway, and inform strategies to improve facility-based postpartum care, strengthen the maternal continuum of care, advance universal health coverage, and support progress toward SDG 3 in Ethiopia[5, 9, 14]. Therefore, this study aimed to determine the prevalence and predictors of missed maternal postpartum assessment before discharge among women who delivered in Ethiopian health facilities using nationally representative 2024–2025 Ethiopia Demographic and Health Survey data.

## Method and Materials

### Study Settings

Ethiopia, located in the Horn of Africa, has an estimated population of about 135 million people in 2025, with most of the population living in rural areas[15]. The country is administratively structured into 12 regional states and two city administrations, which are further divided into zones, woredas, and kebeles. Health services are delivered through a decentralized three-tier health system that includes primary health care units, general hospitals, and specialized referral hospitals. Maternal and newborn health services are provided at different levels of this system, with facility-based childbirth and postpartum care forming key components of the national strategy to reduce maternal and neonatal morbidity and mortality.

### Data Source, Study Population and Sampling Technique

This study analyzed data from the 2024–2025 Ethiopia Demographic and Health Survey (EDHS), a nationally representative survey conducted from August 1, 2024, to April 14, 2025. The EDHS used a two-stage stratified cluster sampling design. First, 805 enumeration areas (281 urban and 524 rural) were selected from the national sampling frame. Then, households were selected systematically within each cluster. Detailed descriptions of the sampling frame, survey procedures, and data collection methods are available in the 2024– 2025 EDHS final report.

The study used data from the Individual Recode (IR) file. The source population included women aged 15–49 years interviewed in the survey who had a recent live birth during the reference period. The study population consisted of women whose most recent live birth occurred in a health facility. Because the outcome variable captured whether a woman received a postpartum assessment before discharge, only institutional births were eligible for inclusion in the analysis. Women with incomplete data on the outcome variable or relevant covariates were excluded. Accordingly, the unit of analysis was women aged 15-49 years with a recent facility-based live birth.

### Study Variables and Measurements

Selection of explanatory variables was informed by previous literature, theoretical relevance, and availability within the EDHS dataset.

### Outcome Variable

The outcome variable was missed postpartum health assessment before discharge among women who gave birth in a health facility. It was categorized as a binary variable: “Yes” for women who did not receive a postpartum health assessment before discharge, coded as 1, and “No” for women who received a postpartum health assessment before discharge, coded as 0.

### Independent Variables

Sociodemographic variables: included maternal age, maternal education, household wealth index, parity, marital status, health insurance status, and perceived birth size (**Figure 1**).

**Figure 1.**
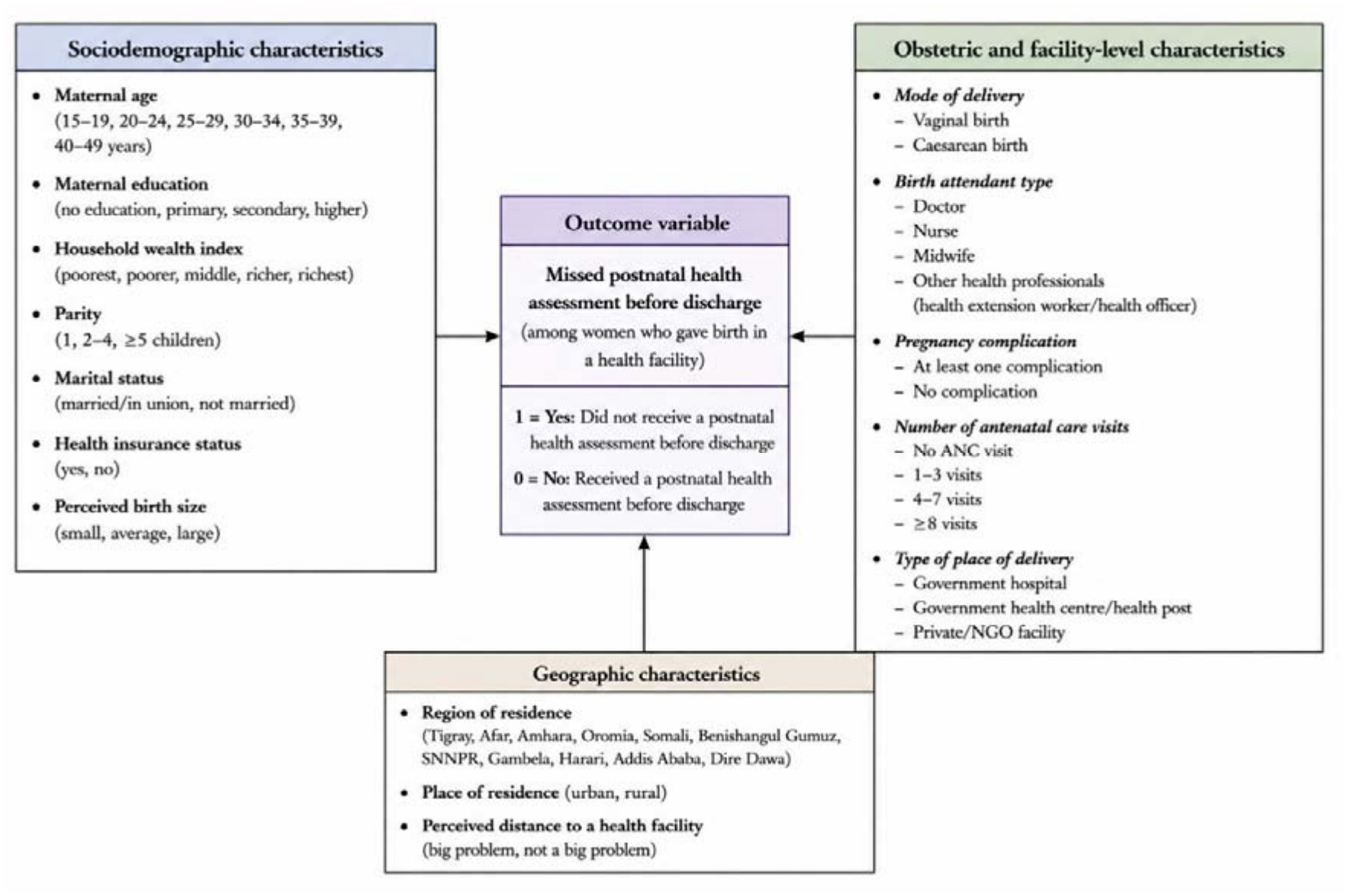
Conceptual framework for the determinants of missed postpartum health assessment before discharge among women who gave birth in health facilities in Ethiopia, EDHS 2024–2025.

Obstetric variables and facility-level variables: included mode of delivery, birth attendant type, pregnancy complication status, and number of antenatal care visits. Mode of delivery was categorized as vaginal or caesarean birth. Birth attendant type was grouped as doctor, nurse, midwife, and other health professionals, which include health extension workers and health officers. Pregnancy complication status was categorized as at least one complication versus none. Antenatal care attendance was categorized as no ANC visit, 1–3 visits, 4–7 visits, and ≥8 visits. Type of place of delivery was categorized as government hospital, government health centre/post, and private/NGO facility.

Geographic variables: Geographic variables included region of residence, place of residence, and perceived distance to a health facility.

## Data Management and Statistical Analysis

Data were extracted from the 2024–2025 Ethiopian Demographic and Health Survey (EDHS), and all data management, recoding, and statistical analyses were performed using Stata version 17. The analysis was restricted to women who gave birth in a health facility, consistent with the study focus on missed postpartum assessment before discharge. The analysis was restricted to 3,645 women who gave birth in a health facility. Sampling weights were subsequently applied to obtain nationally representative estimates. Variables were identified from the EDHS individual recode file, recoded according to the EDHS recode manual, and organized to reflect the study framework. Derived variables were generated where appropriate by recoding and categorizing EDHS variables according to the study objectives. Data completeness was assessed before analysis, and records with missing information for the outcome variable were excluded using a complete-case approach.

Sampling weights were applied throughout the analysis to account for unequal selection probabilities, nonresponse, and the complex survey design, thereby producing nationally interpretable estimates. The survey design variables, including sampling weights, primary sampling units (PSUs), and stratification variables, were incorporated in all descriptive and regression analyses. Because women were nested within enumeration areas, a multilevel mixed-effects logistic regression model was used to account for within-cluster correlation and between-cluster heterogeneity in missed postpartum assessment before discharge. Bivariable multilevel logistic regression analyses were first fitted for each explanatory variable, and variables with p<0.20 were considered for multivariable analysis; variables judged epidemiologically important from previous literature were also retained irrespective of bivariable significance to avoid exclusive reliance on statistical screening.

Four sequential models were specified. Model 0 was an empty model containing only the random intercept. Model I included obstetric and facility-level variables. Model II added sociodemographic variables to Model I. Model III, the final model, added geographic variables to Model II. Because these models were nested, model comparison was based on log-likelihood, deviance, and Wald χ^2^ statistics, and the model with the best overall fit was selected as the final model. Multicollinearity among explanatory variables was assessed using the variance inflation factor, and a mean VIF value of 1.4 indicates the absence of problematic multicollinearity. Fixed effects were reported as adjusted odds ratios (AORs) with 95% confidence intervals, and statistical significance was declared at p<0.05. Random effects were evaluated using the cluster-level variance, intra-class correlation coefficient (ICC), median odds ratio (MOR), and proportional change in variance (PCV) to quantify between-cluster variability and the extent to which successive models explained that variability.

## Results

Of the 3,645 women included in the analysis, the majority delivered at government health centers or posts (58.9%), followed by government hospitals (36.9%) and private/NGO facilities (4.2%). Most women delivered vaginally (87.0%), while 13.0% underwent caesarean section. Nearly half (46.2%) were attended by a midwife, followed by nurses (30.0%), doctors (21.8%), and other health professionals (21.8%).

Slightly over half of the women (51.3%) reported no pregnancy complications, while 48.8% experienced at least one complication. Most women (58.1%) did not have health insurance, compared with 41.9% who were insured. Regarding antenatal care (ANC) attendance, 41.3% had 4–7 visits, 29.0% had ≥8 visits, 20.5% had 1–3 visits, and 9.2% had no ANC visits.

Regarding maternal education, 40.0% had primary education, 36.2% had secondary or higher education, and 23.7% had no formal education. By household wealth, the largest proportion of women were in the richest quintile (33.1%), followed by richer (26.4%), middle (17.5%), poorer (13.6%), and poorest (9.4%) quintiles.

Most participants were aged 20–29 years (54.9%), followed by 30–39 years (35.9%), with smaller proportions aged ≤19 years (4.1%) and 40–49 years (5.2%). By parity, over half were multiparous (54.3%), while 27.8% were primiparous and 17.9% were grand multiparous. The vast majority of women were currently married (93.4%), with small proportions formerly married (5.6%) or never married (1.0%).

Regarding perceived baby birth size, nearly half (47.3%) perceived their baby as average size, 20.0% as larger than average, 14.0% as very large, 12.6% as very small, and 6.2% as smaller than average. Slightly more than half of the women resided in rural areas (56.1%) compared with urban areas (43.9%). Most women did not consider distance to a health facility a big problem (69.2%), while 30.8% did.

Geographically, the largest proportions of women were from Oromia (34.1%) and Amhara (28.8%), followed by Addis Ababa (7.2%), South Ethiopia (6.3%), and Tigray (6.6%), with the remaining regions each accounting for less than 6% of the sample (**Table 1**).

**Table 1.**
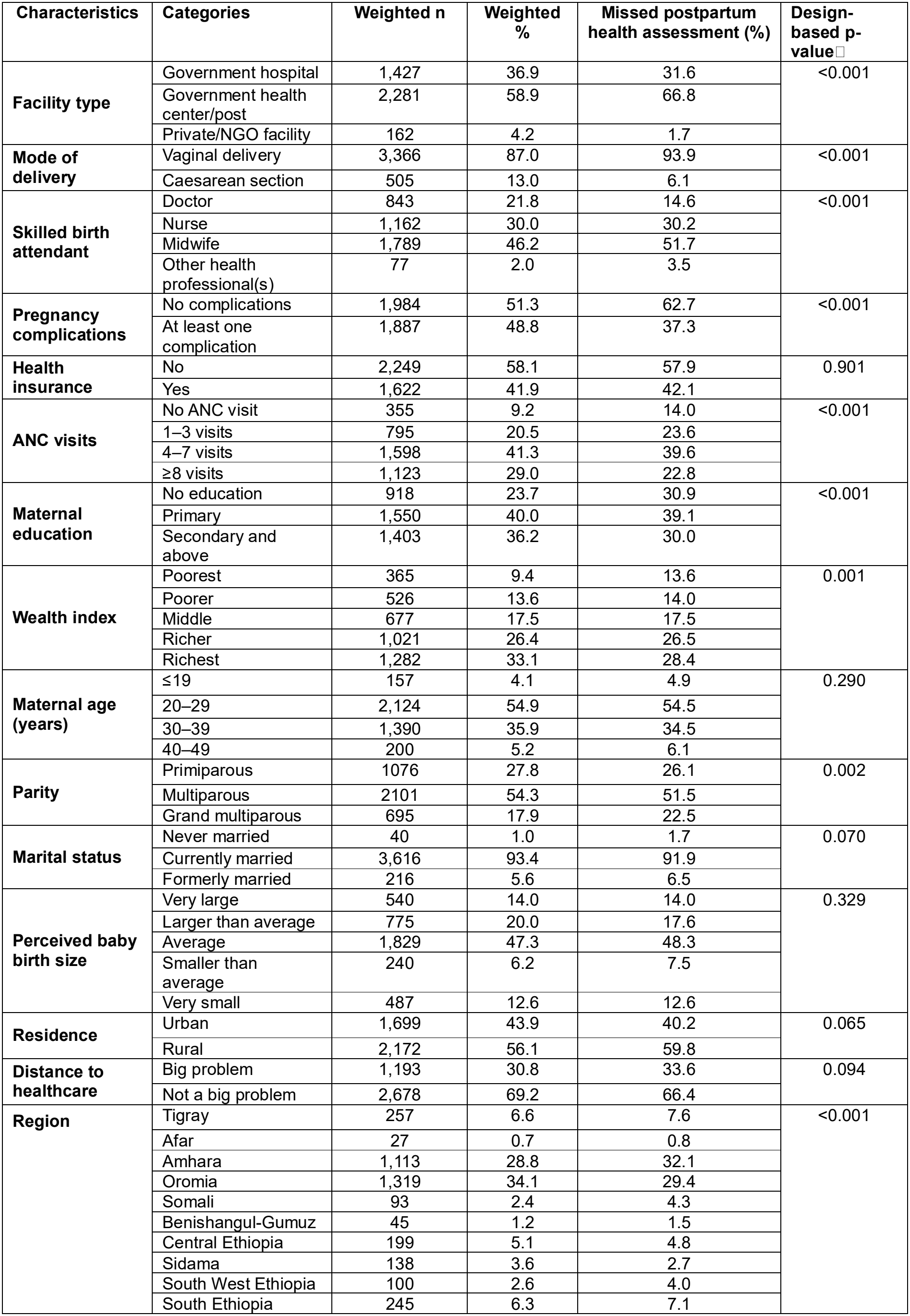
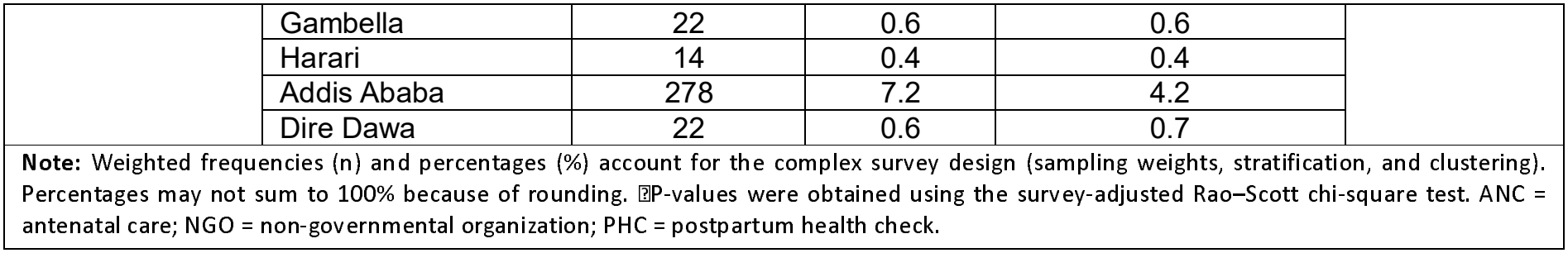
Weighted distribution of maternal, obstetric, and health service characteristics by missed postpartum health assessment before discharge among women who gave birth in Ethiopia (n = 3,645).

| Characteristics | Categories | Weighted n | Weighted % | Missed postpartum health assessment (%) | Design-based p-value□ |
|---|---|---|---|---|---|
| **Facility type** | Government hospital | 1,427 | 36.9 | 31.6 | <0.001 |
|  | Government health center/post | 2,281 | 58.9 | 66.8 |  |
|  | Private/NGO facility | 162 | 4.2 | 1.7 |  |
| **Mode of delivery** | Vaginal delivery | 3,366 | 87.0 | 93.9 | <0.001 |
|  | Caesarean section | 505 | 13.0 | 6.1 |  |
| **Skilled birth attendant** | Doctor | 843 | 21.8 | 14.6 | <0.001 |
|  | Nurse | 1,162 | 30.0 | 30.2 |  |
|  | Midwife | 1,789 | 46.2 | 51.7 |  |
|  | Other health professional(s) | 77 | 2.0 | 3.5 |  |
| **Pregnancy complications** | No complications | 1,984 | 51.3 | 62.7 | <0.001 |
|  | At least one complication | 1,887 | 48.8 | 37.3 |  |
| **Health insurance** | No | 2,249 | 58.1 | 57.9 | 0.901 |
|  | Yes | 1,622 | 41.9 | 42.1 |  |
| **ANC visits** | No ANC visit | 355 | 9.2 | 14.0 | <0.001 |
|  | 1–3 visits | 795 | 20.5 | 23.6 |  |
|  | 4–7 visits | 1,598 | 41.3 | 39.6 |  |
|  | ≥8 visits | 1,123 | 29.0 | 22.8 |  |
| **Maternal education** | No education | 918 | 23.7 | 30.9 | <0.001 |
|  | Primary | 1,550 | 40.0 | 39.1 |  |
|  | Secondary and above | 1,403 | 36.2 | 30.0 |  |
| **Wealth index** | Poorest | 365 | 9.4 | 13.6 | 0.001 |
|  | Poorer | 526 | 13.6 | 14.0 |  |
|  | Middle | 677 | 17.5 | 17.5 |  |
|  | Richer | 1,021 | 26.4 | 26.5 |  |
|  | Richest | 1,282 | 33.1 | 28.4 |  |
| **Maternal age (years)** | ≤19 | 157 | 4.1 | 4.9 | 0.290 |
|  | 20–29 | 2,124 | 54.9 | 54.5 |  |
|  | 30–39 | 1,390 | 35.9 | 34.5 |  |
|  | 40–49 | 200 | 5.2 | 6.1 |  |
| **Parity** | Primiparous | 1076 | 27.8 | 26.1 | 0.002 |
|  | Multiparous | 2101 | 54.3 | 51.5 |  |
|  | Grand multiparous | 695 | 17.9 | 22.5 |  |
| **Marital status** | Never married | 40 | 1.0 | 1.7 | 0.070 |
|  | Currently married | 3,616 | 93.4 | 91.9 |  |
|  | Formerly married | 216 | 5.6 | 6.5 |  |
| **Perceived baby birth size** | Very large | 540 | 14.0 | 14.0 | 0.329 |
|  | Larger than average | 775 | 20.0 | 17.6 |  |
|  | Average | 1,829 | 47.3 | 48.3 |  |
|  | Smaller than average | 240 | 6.2 | 7.5 |  |
|  | Very small | 487 | 12.6 | 12.6 |  |
| **Residence** | Urban | 1,699 | 43.9 | 40.2 | 0.065 |
|  | Rural | 2,172 | 56.1 | 59.8 |  |
| **Distance to healthcare** | Big problem | 1,193 | 30.8 | 33.6 | 0.094 |
|  | Not a big problem | 2,678 | 69.2 | 66.4 |  |
| **Region** | Tigray | 257 | 6.6 | 7.6 | <0.001 |
|  | Afar | 27 | 0.7 | 0.8 |  |
|  | Amhara | 1,113 | 28.8 | 32.1 |  |
|  | Oromia | 1,319 | 34.1 | 29.4 |  |
|  | Somali | 93 | 2.4 | 4.3 |  |
|  | Benishangul-Gumuz | 45 | 1.2 | 1.5 |  |
|  | Central Ethiopia | 199 | 5.1 | 4.8 |  |
|  | Sidama | 138 | 3.6 | 2.7 |  |
|  | South West Ethiopia | 100 | 2.6 | 4.0 |  |
|  | South Ethiopia | 245 | 6.3 | 7.1 |  |
|  | Gambella | 22 | 0.6 | 0.6 |  |
|  | Harari | 14 | 0.4 | 0.4 |  |
|  | Addis Ababa | 278 | 7.2 | 4.2 |  |
|  | Dire Dawa | 22 | 0.6 | 0.7 |  |

**Note:** Weighted frequencies (n) and percentages (%) account for the complex survey design (sampling weights, stratification, and clustering). Percentages may not sum to 100% because of rounding. □P-values were obtained using the survey-adjusted Rao–Scott chi-square test. ANC = antenatal care; NGO = non-governmental organization; PHC = postpartum health check.

## Postpartum Health Assessment Before Discharge Status in Ethiopia

A total of 3,645 women who delivered at a health facility were included in the analysis. Overall, 40.1% (95% CI: 37.0–43.3%) of women did not receive a postpartum health assessment before discharge (**Figure 2**).

**Figure 2.**
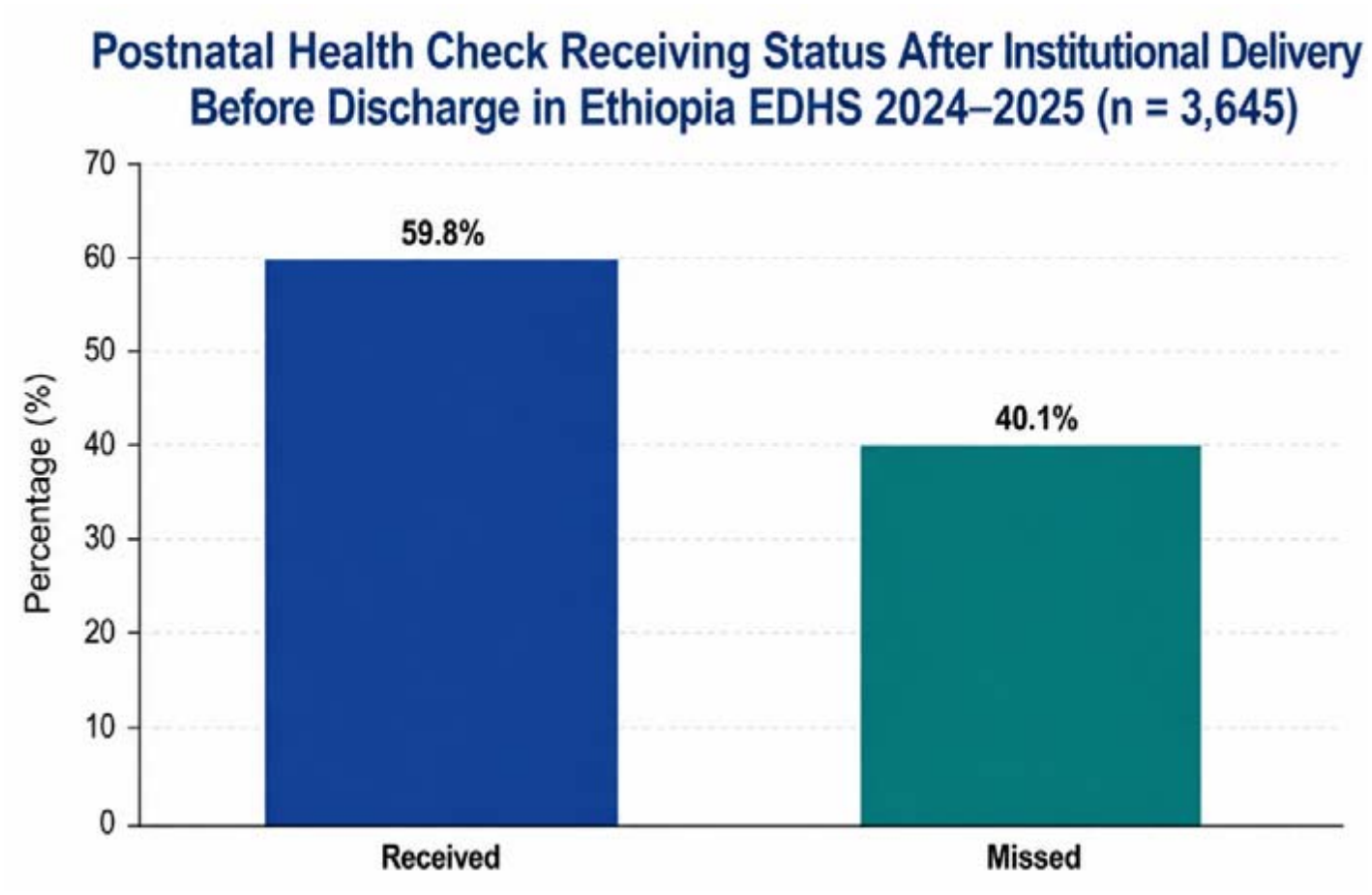
Weighted prevalence of postpartum health check status before discharge among women who gave birth in health facilities in Ethiopia, EDHS 2024–2025.

## Determinants of Postpartum Health Assessment Before Discharge

### Random Effects Analysis

The null model revealed substantial between-cluster variation in missed postpartum assessment before discharge, with a cluster-level variance of 0.945 and an intraclass correlation coefficient (ICC) of 22.3%, indicating that approximately one-fifth of the total variability in the outcome was attributable to differences between clusters rather than individual-level characteristics. This finding justified the use of a multilevel modelling approach.

Introducing obstetric and facility-level variables (Model I) reduced the cluster variance to 0.900 and the ICC to 21.5%, corresponding to a proportional change in variance (PCV) of 4.8% relative to the null model, indicating that these factors explained a modest share of the observed clustering. The subsequent addition of sociodemographic variables (Model II) did not yield a further sequential reduction in cluster-level variance; instead, the cluster variance rose to 0.941 and the ICC to 22.2%, values close to those of the null model, corresponding to a net PCV of only 0.4% relative to the null model. This pattern suggests that sociodemographic characteristics accounted for little additional between-cluster heterogeneity once obstetric and facility-level factors were held constant, and may partly offset the variance reduction achieved in Model I, potentially reflecting compositional overlap between sociodemographic characteristics and obstetric or facility-level factors already captured in the model. Fixed-effects model fit nonetheless continued to improve at this stage, with the log pseudolikelihood increasing from −2,261.38 (Model I) to −2,228.86 (Model II) and the Wald χ^2^ statistic rising from 117.48 to 143.93, indicating that sociodemographic variables contributed to explaining individual-level variation in the outcome despite their limited contribution to cluster-level heterogeneity.

The final model (Model III), which additionally incorporated geographic variables, produced the largest reduction in unexplained between-cluster variation, lowering the cluster variance to 0.811 and the ICC to 19.8% (PCV = 14.2% relative to the null model). Consistent with this trend, the median odds ratio (MOR) declined from 2.53 in the null model to 2.47 in Model I, rose to 2.52 in Model II, and declined further to 2.36 in the final model — a pattern that mirrors the PCV and ICC trends and indicates that geographic context, rather than individual sociodemographic characteristics, accounted for the majority of the explainable between-cluster heterogeneity in missed postpartum assessment. Model fit improved progressively and significantly across the full model-building sequence, with the log pseudolikelihood increasing from −2,435.08 (null model) to −2,202.67 (Model III) and the Wald χ^2^ statistic rising to 220.70 in the final model, confirming that each successive model provided a significantly better fit to the data than the null model. Based on these comparative fit statistics, Model III — incorporating obstetric, facility-level, sociodemographic, and geographic variables — was selected as the final model for identifying factors associated with missed postpartum assessment before discharge (**Table 2**).

**Table 2.** Random-effects parameters and model fit statistics for the multilevel logistic regression models predicting missed postpartum assessment before discharge, Ethiopia, 2024–2025 (n = 3,645)

| **Measure** | **Null Model** | **Model I** | **Model II** | **Model III** |
|---|---|---|---|---|
| **Cluster variance (SE)** | 0.945 | 0.900 | 0.941 | 0.811 |
| **ICC (%)** | 22.3 | 21.5 | 22.2 | 19.8 |
| **MOR** | 2.53 | 2.47 | 2.52 | 2.36 |
| **PCV (%)** | Reference | 4.8 | 0.4 | 14.2 |
| **Log pseudolikelihood** | −2435.08 | −2261.38 | −2228.86 | −2202.67 |
| **Wald χ2** | — | 117.48 | 143.93 | 220.70 |

*ICC, intraclass correlation coefficient; MOR, median odds ratio; PCV, proportional change in variance (relative to the Null Model). Model I: obstetric and facility-level variables. Model II: Model I + socio-demographic variables. Model III (final model): Model II + geographic variables. Wald χ² tests the joint significance of fixed effects added at each stage relative to the Null Model.*

**Table 3.**
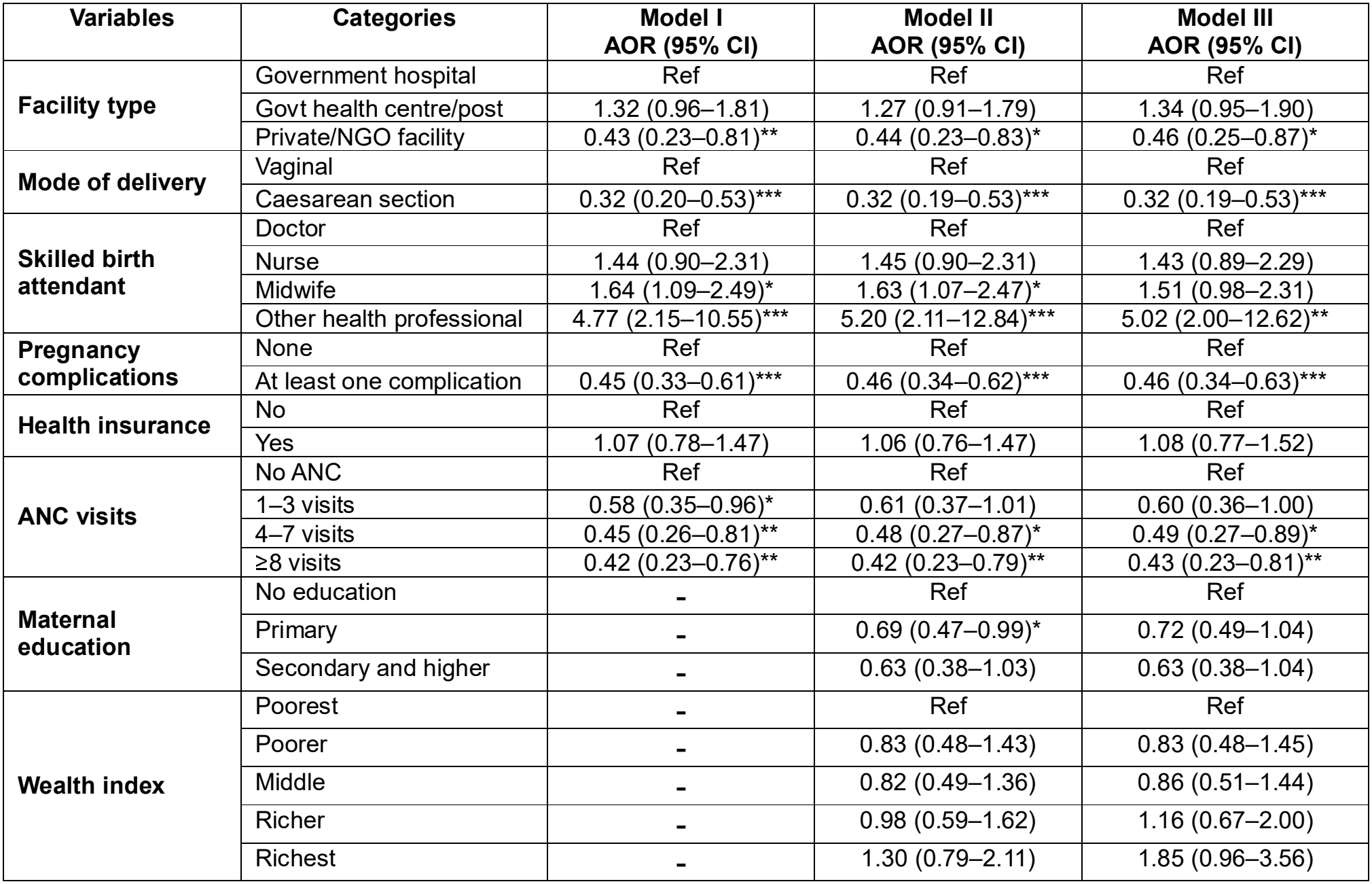
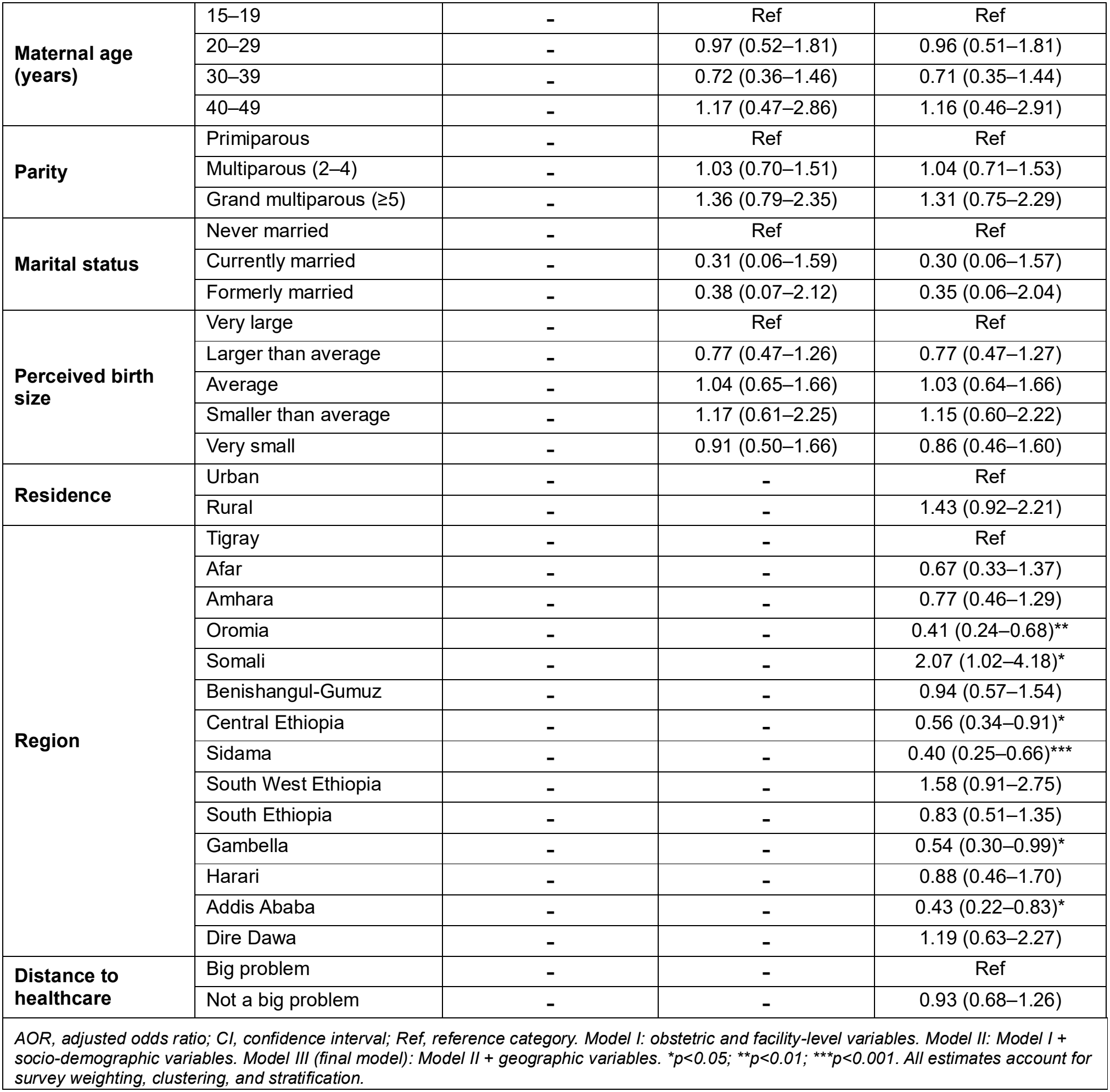
Hierarchical multivariable logistic regression of predictors of missed postpartum assessment before discharge, Ethiopia, 2024–2025 (n = 3,645)

| **Variables** | **Categories** | **Model I<br>AOR (95% CI)** | **Model II<br>AOR (95% CI)** | **Model III<br>AOR (95% CI)** |
|---|---|---|---|---|
| **Facility type** | Government hospital | Ref | Ref | Ref |
|  | Govt health centre/post | 1.32 (0.96–1.81) | 1.27 (0.91–1.79) | 1.34 (0.95–1.90) |
|  | Private/NGO facility | 0.43 (0.23–0.81)** | 0.44 (0.23–0.83)* | 0.46 (0.25–0.87)* |
| **Mode of delivery** | Vaginal | Ref | Ref | Ref |
|  | Caesarean section | 0.32 (0.20–0.53)*** | 0.32 (0.19–0.53)*** | 0.32 (0.19–0.53)*** |
| **Skilled birth attendant** | Doctor | Ref | Ref | Ref |
|  | Nurse | 1.44 (0.90–2.31) | 1.45 (0.90–2.31) | 1.43 (0.89–2.29) |
|  | Midwife | 1.64 (1.09–2.49)* | 1.63 (1.07–2.47)* | 1.51 (0.98–2.31) |
|  | Other health professional | 4.77 (2.15–10.55)*** | 5.20 (2.11–12.84)*** | 5.02 (2.00–12.62)** |
| **Pregnancy complications** | None | Ref | Ref | Ref |
|  | At least one complication | 0.45 (0.33–0.61)*** | 0.46 (0.34–0.62)*** | 0.46 (0.34–0.63)*** |
| **Health insurance** | No | Ref | Ref | Ref |
|  | Yes | 1.07 (0.78–1.47) | 1.06 (0.76–1.47) | 1.08 (0.77–1.52) |
| **ANC visits** | No ANC | Ref | Ref | Ref |
|  | 1–3 visits | 0.58 (0.35–0.96)* | 0.61 (0.37–1.01) | 0.60 (0.36–1.00) |
|  | 4–7 visits | 0.45 (0.26–0.81)** | 0.48 (0.27–0.87)* | 0.49 (0.27–0.89)* |
|  | ≥8 visits | 0.42 (0.23–0.76)** | 0.42 (0.23–0.79)** | 0.43 (0.23–0.81)** |
| **Maternal education** | No education | - | Ref | Ref |
|  | Primary | - | 0.69 (0.47–0.99)* | 0.72 (0.49–1.04) |
|  | Secondary and higher | - | 0.63 (0.38–1.03) | 0.63 (0.38–1.04) |
| **Wealth index** | Poorest | - | Ref | Ref |
|  | Poorer | - | 0.83 (0.48–1.43) | 0.83 (0.48–1.45) |
|  | Middle | - | 0.82 (0.49–1.36) | 0.86 (0.51–1.44) |
|  | Richer | - | 0.98 (0.59–1.62) | 1.16 (0.67–2.00) |
|  | Richest | - | 1.30 (0.79–2.11) | 1.85 (0.96–3.56) |
| **Maternal age (years)** | 15–19 | - | Ref | Ref |
|  | 20–29 | - | 0.97 (0.52–1.81) | 0.96 (0.51–1.81) |
|  | 30–39 | - | 0.72 (0.36–1.46) | 0.71 (0.35–1.44) |
|  | 40–49 | - | 1.17 (0.47–2.86) | 1.16 (0.46–2.91) |
| **Parity** | Primiparous | - | Ref | Ref |
|  | Multiparous (2–4) | - | 1.03 (0.70–1.51) | 1.04 (0.71–1.53) |
|  | Grand multiparous (≥5) | - | 1.36 (0.79–2.35) | 1.31 (0.75–2.29) |
| **Marital status** | Never married | - | Ref | Ref |
|  | Currently married | - | 0.31 (0.06–1.59) | 0.30 (0.06–1.57) |
|  | Formerly married | - | 0.38 (0.07–2.12) | 0.35 (0.06–2.04) |
| **Perceived birth size** | Very large | - | Ref | Ref |
|  | Larger than average | - | 0.77 (0.47–1.26) | 0.77 (0.47–1.27) |
|  | Average | - | 1.04 (0.65–1.66) | 1.03 (0.64–1.66) |
|  | Smaller than average | - | 1.17 (0.61–2.25) | 1.15 (0.60–2.22) |
|  | Very small | - | 0.91 (0.50–1.66) | 0.86 (0.46–1.60) |
| **Residence** | Urban | - | - | Ref |
|  | Rural | - | - | 1.43 (0.92–2.21) |
| **Region** | Tigray | - | - | Ref |
|  | Afar | - | - | 0.67 (0.33–1.37) |
|  | Amhara | - | - | 0.77 (0.46–1.29) |
|  | Oromia | - | - | 0.41 (0.24–0.68)** |
|  | Somali | - | - | 2.07 (1.02–4.18)* |
|  | Benishangul-Gumuz | - | - | 0.94 (0.57–1.54) |
|  | Central Ethiopia | - | - | 0.56 (0.34–0.91)* |
|  | Sidama | - | - | 0.40 (0.25–0.66)*** |
|  | South West Ethiopia | - | - | 1.58 (0.91–2.75) |
|  | South Ethiopia | - | - | 0.83 (0.51–1.35) |
|  | Gambella | - | - | 0.54 (0.30–0.99)* |
|  | Harari | - | - | 0.88 (0.46–1.70) |
|  | Addis Ababa | - | - | 0.43 (0.22–0.83)* |
|  | Dire Dawa | - | - | 1.19 (0.63–2.27) |
| **Distance to healthcare** | Big problem | - | - | Ref |
|  | Not a big problem | - | - | 0.93 (0.68–1.26) |

*AOR, adjusted odds ratio; CI, confidence interval; Ref, reference category. Model I: obstetric and facility-level variables. Model II: Model I + socio-demographic variables. Model III (final model): Model II + geographic variables. \*p<0.05; \*\*p<0.01; \*\*\*p<0.001. All estimates account for survey weighting, clustering, and stratification.*

### Fixed Effects Analysis

In the final multivariable model (Model III), which adjusted for obstetric, facility-level, sociodemographic, and geographic factors, several variables were independently associated with missed postpartum assessment.

Facility-level and obstetric factors: Compared with women who delivered in government hospitals, those who delivered in private/NGO facilities had significantly lower odds of missed postpartum assessment (AOR 0.46, 95% CI 0.25–0.87), while odds did not differ significantly for government health centres/posts (AOR 1.34, 95% CI 0.95–1.90). Caesarean delivery was associated with substantially lower odds of missed assessment compared with vaginal delivery (AOR 0.32, 95% CI 0.19–0.53). Relative to doctor-attended births, deliveries attended by other health professionals had markedly higher odds of missed assessment (AOR 5.02, 95% CI 2.00–12.62), and midwife-attended births (AOR 1.51, 95% CI 0.98–2.31) did not differ significantly from doctor-attended births. Women with at least one pregnancy complication had significantly lower odds of missed assessment than those without complications (AOR 0.46, 95% CI 0.34–0.63). A dose–response relationship was observed for antenatal care attendance, with significantly lower odds of missed assessment among women with 4–7 ANC visits (AOR 0.49, 95% CI 0.27–0.89) and ≥8 ANC visits (AOR 0.43, 95% CI 0.23–0.81) compared with women with no ANC visits; the association for 1–3 visits did not reach statistical significance (AOR 0.60, 95% CI 0.36–1.00).

Sociodemographic factors: Health insurance status, maternal education, wealth index, maternal age, parity, marital status, and perceived birth size were not significantly associated with missed postpartum assessment after adjustment for geographic factors in the final model.

Geographic factors: Region of residence remained significantly associated with missed postpartum assessment after full adjustment, compared with Tigray, women residing in Oromia (AOR 0.41, 95% CI 0.24–0.68), Sidama (AOR 0.40, 95% CI 0.25–0.66), Central

Ethiopia (AOR 0.56, 95% CI 0.34–0.91), Gambella (AOR 0.54, 95% CI 0.30–0.99), and Addis Ababa (AOR 0.43, 95% CI 0.22–0.83) had significantly lower odds of missed assessment, while women in the Somali region had significantly higher odds (AOR 2.07, 95% CI 1.02–4.18). No other regions differed significantly from Tigray. Rural residence (AOR 1.43, 95% CI 0.92–2.21) and perceiving distance to a health facility as “not a big problem” (AOR 0.93, 95% CI 0.68–1.26) were not significantly associated with the outcome.

## Discussion

Our study shows that 40.1% of women delivering in Ethiopian health facilities did not receive a postpartum assessment before discharge. This appears high from a quality-of-care perspective, although directly comparable Ethiopian estimates of missed pre-discharge maternal assessment are not available; prior Ethiopian studies and DHS-based analyses have instead measured postpartum care use or check-ups within 48 hours after birth, not whether an assessment occurred before leaving the facility[9, 16, 17].

In Ethiopia, earlier nationally representative analyses reported very low maternal postpartum care coverage within two days of birth, including 17% in the 2016 EDHS-based analysis and 6.9% in another DHS analysis using a broader denominator of recent births[16, 18, 19]. Among women delivering in facilities, one Ethiopian multilevel study reported that 42% received postpartum care within two days, and also noted that most facility-born women did not receive proper postpartum care and that over two-thirds of vaginal births were discharged within 24 hours[9]. At the subnational level, quality-adjusted newborn postpartum care coverage in Ethiopia was only 9% in a 2016 EDHS analysis, and a later study using the 2019 mini-EDHS found a large gap between newborn postpartum contact (34.8%) and effective coverage (10.4%)[20, 21]. Taken together, these findings suggest that the present estimate is not directly comparable with prior Ethiopian postpartum utilization metrics, but it is consistent with a broader pattern in which service contact and effective postpartum care content remain substantially misaligned[20, 21].

This study therefore makes a distinct contribution by moving beyond conventional postpartum care utilization indicators to examine whether women actually received an assessment before discharge, which is better interpreted as an indicator of intrafacility maternal care quality than of service contact alone[5]. Unlike previous Ethiopian studies that focused on postpartum care attendance after childbirth or within two days of birth, this analysis isolates a critical but under-measured transition point: whether facility birth culminated in a documented maternal assessment before the woman left care[9, 17]. That distinction is important because WHO guidance places explicit emphasis on discharge readiness, quality of care, and assessment before women and newborns leave the facility, while recent measurement reviews have shown that several recommended pre-discharge criteria remain poorly captured in routine monitoring systems[7]. Methodologically, the use of the 2024–2025 EDHS, a nationally weighted sample, and a multilevel mixed-effects model incorporating individual, obstetric, facility, and regional factors strengthens the study’s novelty and policy relevance for contemporary Ethiopia[17].

Comparable evidence from sub-Saharan Africa also suggests that the level observed here is concerning. A multi-country DHS analysis cited in the Ghana facility study estimated that only 66.6% of women across 33 sub-Saharan African countries received a pre-discharge check by a health professional, implying that roughly one-third did not, with marked cross-country variation from 26.6% in Eswatini to 94.4% in Burkina Faso[22]. In a broader meta-analysis across 36 sub-Saharan African countries, pooled postpartum care utilization was 52.5%, while Ethiopia was among the lower-coverage settings in the regional comparison[9]. A 2025 systematic review across LMICs similarly estimated pooled postpartum care utilization at 48.9%, again indicating that postpartum care remains less consistently delivered than antenatal or delivery care in many settings[23]. Even so, these cross-country figures should be interpreted cautiously because most refer to postpartum care utilization or postpartum checks, not specifically to missed maternal assessment before discharge, which reinforces the novelty of the present estimate rather than allowing direct one-to-one comparison[7, 22].

The lower odds of missed assessment among women delivering in private or NGO facilities than in government hospitals likely indicate differences in care processes rather than ownership alone. A possible explanation is that private and NGO facilities may have lower caseloads, shorter queues, or more tightly supervised discharge procedures, whereas government hospitals may face heavier referral burdens and more complex workflows; however, this interpretation should remain cautious because ownership may also proxy urban location, case-mix, or socioeconomic selection that were not fully measured. This interpretation is broadly consistent with Ethiopian evidence showing that private facilities were associated with higher-quality PNC than public facilities in national DHS-based analysis, although that study assessed composite PNC quality rather than maternal assessment before discharge[17].

The strong association with caesarean birth is also consistent with prior Ethiopian and sub-Saharan literature on postpartum care use[9, 16, 24]. One plausible explanation is that women undergoing caesarean section remain in facilities longer and have more postoperative reviews, which creates more opportunities for formal postpartum assessment before discharge, rather than the operation itself intrinsically improving care quality[9]. By contrast, women with uncomplicated vaginal births may be discharged rapidly and treated as lower risk, despite evidence that major postpartum complications can still emerge after apparently normal delivery [25]. The implication is that universal postpartum assessment is needed, rather than a model in which formal review is triggered mainly by surgical delivery or overt complications.

The finding that births attended by other health professionals (extension workers or health officers) had higher odds of missed assessment than doctor-attended births should likewise be interpreted as a signal of how care is organised, not of professional worth. A plausible explanation is that gaps in handover, documentation, or role clarity around who is responsible for the discharge assessment may contribute to missed checks when care pathways are fragmented. This aligns with broader evidence that quality failures in postpartum care often reflect weakly specified processes of assessment and transition, and that women in sub-Saharan Africa frequently report compromised quality, poor communication, and limited accountability in facility maternity care[26].

Women reporting pregnancy complications were less likely to leave without assessment, which appears to reflect risk-responsive clinical attention. However, the converse finding is equally important: women without recognised complications may be systematically overlooked, even though severe postpartum morbidity can evolve rapidly after an uncomplicated labour. This again supports a universal approach to immediate postpartum assessment, with risk stratification used to determine intensity of follow-up rather than whether assessment occurs at all[5].

The graded association between ANC attendance and lower odds of missed postpartum assessment is one of the most policy-relevant findings. Prior Ethiopian studies consistently found that four or more ANC visits were associated with higher postpartum care use, and national and subnational analyses have similarly linked ANC to postpartum care utilization and quality-adjusted care along the continuum[18, 21, 23, 27]. One plausible explanation is that ANC increases counselling, preparedness, familiarity with facilities, and women’s expectations regarding postpartum checks, but the current data do not directly test these mechanisms[21]. Even so, the consistency of this pattern supports using ANC not only to promote facility delivery, but also to prepare women for immediate postpartum assessment before discharge[22, 28].

The residual regional variation after adjustment indicates that missed postpartum assessment is also shaped by subnational health-system differences. This is consistent with Ethiopian evidence showing substantial geographic clustering and regional inequalities in postpartum care use, maternal continuum-of-care dropout, and quality-adjusted newborn postpartum care, with Somali repeatedly emerging as a higher-risk region in prior work[18, 20, 29]. These differences may reflect variation in readiness, workforce distribution, programme implementation strength, and fragility, but those pathways were not directly measured in the current survey and should therefore be interpreted as plausible system-level explanations rather than established causal mechanisms[30].

A notable implication of the null associations for several socioeconomic variables after adjustment is that, once women have already reached a facility for childbirth, the dominant failures may shift from access barriers to within-facility quality gaps. This interpretation is compatible with broader Ethiopian and LMIC evidence showing that social disadvantage remains strongly associated with postpartum care utilization overall, while effective coverage analyses increasingly show that contact does not ensure receipt of essential care content[20, 23].

## Strength and Limitation

This study has several important strengths. First, it used a nationally weighted sample of 3,645 facility births from the 2024–2025 Ethiopia Demographic and Health Survey, which enhances the national representativeness and contemporary policy relevance of the findings. Second, the analysis focused on an underexamined but programmatically important outcome: whether women received a postpartum assessment before discharge after institutional birth, thereby capturing an aspect of intrafacility quality of care rather than service contact alone. Third, the use of a multilevel mixed-effects model accounted for both individual- and cluster-level heterogeneity and incorporated survey weighting, clustering, and stratification, which improved the robustness of the estimates and strengthened the validity of the inferences drawn at national level.

Several limitations should also be considered when interpreting these findings. First, the cross-sectional nature of the survey precludes causal inference, and the observed associations should therefore be interpreted as correlational rather than causal. Second, the outcome measure captured only whether any postpartum assessment occurred before discharge and did not provide information on the timing, completeness, technical content, or adherence to standard discharge criteria, which limits conclusions about the quality of the assessment itself. Third, as a secondary analysis of survey data, the study was unable to account for potentially important facility-level determinants, including staffing levels, caseload, provider workload, supervision, and local discharge practices, all of which may influence whether essential postpartum checks are completed before women leave the facility

## Recommendation

These findings support a more operational set of recommendations than increasing institutional delivery alone. Ethiopia should consider making documented maternal postpartum assessment before discharge a routine standard for all facility births, supported by standardized discharge checklists, postpartum observation protocols for both vaginal and caesarean births, and explicit alignment with WHO postpartum care recommendations on quality, discharge readiness, and continuity of care. Given the association with facility type and provider cadre, government hospitals in particular may benefit from competency-based in-service training, clearer task allocation, stronger provider accountability, and routine audit- and-feedback systems that verify whether essential discharge assessments were completed and recorded. At the health information level, inclusion of pre-discharge postpartum assessment indicators in Ethiopia’s HMIS and electronic discharge documentation could improve routine measurement, especially given the documented gaps in existing postpartum quality metrics and the need for better monitoring of provision-of-care processes. Regional heterogeneity also argues for targeted quality-improvement support in high-burden settings, particularly underserved regions such as Somali, rather than reliance on uniform national strategies. Finally, because ANC attendance was strongly associated with lower odds of missed assessment, immediate postpartum assessment counselling and discharge expectations should be integrated explicitly into routine ANC communication so that women know a facility birth should end not simply with delivery, but with a completed maternal check before going home.

## Data Availability

Data are available in a public, open-access repository. Data are available online and can be accessed at https://dhsprogram.com/data/available-datasets.cfm.

## Declarations

### Ethical Approval and Consent to Participate

Since the study was a secondary data analysis of publicly available survey data from the DHS program, ethical approval and participant consent were not necessary for this particular study. We requested DHS Program and permission was granted to download and use the data for this study. The DHS Program adheres to rigorous ethical standards reviewed and approved by both the host country’s institutional review board and the ICF International Institutional Review Board. All materials used in this study have been appropriately acknowledged in compliance with DHS protocols. This study used secondary data analysis, hence no direct participation from individuals was required. This study used exclusively de-identified information, ensuring full compliance with ethical standards for participant privacy and confidentiality.

### Consent for publication

Not applicable.

### Authors’ contributions

Conceptualization: BAK.

Methodology: BAK, EAT.

Data curation: BAK.

Formal analysis: BAK.

Investigation: SAB, DMM, BYM.

Validation: BAK, EAT, SAB, DMM, BYM.

Writing - original draft: BAK.

Writing - review and editing: BAK, EAT, SAB, DMM, BYM.

Supervision: EAT.

All authors have read and approved the final manuscript.

## Abbreviations

AOR: Adjusted odds ratio
CI: Confidence interval
DHS: Demographic and Health Survey
EDHS: Ethiopia Demographic and Health Survey
HMIS: Health Management Information System
ICC: Intraclass correlation coefficient
IR: Individual Recode (file)
LMIC(s): Low- and middle-income countries
MOR: Median odds ratio
NGO: Non-governmental organization
PCV: Proportional change in variance
PHC: Postpartum health check
PSU: Primary sampling unit
SDG: Sustainable Development Goal
VIF: Variance inflation factor
WHO: World Health Organization

